# Ecological and molecular drivers of ESBL plasmid dissemination in *Enterobacteriaceae* in Vietnam

**DOI:** 10.64898/2026.05.06.26352499

**Authors:** Phuong Pham, Pham Thi Mong Quynh, Quynh Nguyen, Ha Thanh Tuyen, Nguyen Thi Nguyen To, Le Thi Quynh Nhi, Vu Thuy Duong, Nguyen Phu Huong Lan, Stephen Baker, Guy Thwaites, Maia A. Rabaa, Hao Chung The, Christoph M. Tang, Pham Thanh Duy

**Author notes:** Equal Contribution (Christoph M. Tang, Pham Thanh Duy). Correspondence: Dr. Pham Thanh Duy, Molecular Epidemiology Group, Oxford University Clinical Research Unit, Vietnam. 764 Vo Van Kiet Street, Ho Chi Minh City, Vietnam;.

## Abstract

**Background:** Extended-spectrum beta-lactamase (ESBL)-producing *Enterobacteriaceae* are among the WHO’s highest-priority antibiotic-resistant pathogens. Plasmids are the main drivers of ESBL dissemination, yet their reservoirs and transmission dynamics remain poorly understood in low- and middle-income countries such as Vietnam, where infections caused by ESBL-producing bacteria are prevalent.

**Methods:** Here, we characterised the genetic structure of 68 ESBL-encoding conjugative plasmids isolated from the human gut microbiome (HGM) of healthy Vietnamese children. We further examined the extent of ESBL plasmid transfer between the HGM and human disease-causing *Enterobacteriaceae* pathogens (including *Shigella sonnei*, non-typhoidal *Salmonella*, extraintestinal pathogenic *Escherichia coli* ST131), as well as *E. coli* isolated from animals.

**Results:** The dominant plasmid Inc groups found in cephalosporin-resistant human gut bacteria were IncF, IncB/O/K/Z and IncI1, carrying mainly *bla*_*CTX-M-14*_, *bla*_*CTX-M-15*_, *bla*_*CTX-M-27*_ and *bla*_*CTX-M-55*_. These plasmids from the HGM, rather than from animal *E. coli*, share higher genetic similarity to plasmids in human pathogens, suggesting that human gut is the main reservoir for clinically relevant ESBL plasmids. We also found that widespread ESBL plasmid variants exhibited higher conjugation frequencies, facilitating broader geographical and host dissemination. In contrast, less mobile plasmids persisted mainly through clonal expansion of their bacterial hosts.

**Conclusions:** These findings highlight the central role of the human gut as a reservoir for ESBL plasmids and provide insights into the biological factors contributing to their successful spread among pathogenic *Enterobacteriaceae*. Our work underscores the need for targeted interventions to reduce colonization and transmission of ESBL-producing bacteria within the human gut.

## Background

Antimicrobial resistance (AMR) is a global threat, with estimates projecting >10 million deaths annually by 2050^1^. Therefore, it is critical to understand the mechanisms of AMR dissemination between bacteria to devise approaches to limit its emergence and spread. Bacterial conjugation involves the transfer of plasmids through direct cell-to-cell contact and is considered the most widespread mechanism for the dissemination of AMR^2^. In particular, resistance to clinically important β-lactams, particularly third-generation cephalosporins mediated by the production of extended-spectrum beta lactamases (ESBL), poses a significant public health concern, as ESBL-encoding plasmids can spread horizontally by conjugation^3^.

The intestinal tract of humans and animals is a recognised “hotspot” for AMR genes, where the high density and diversity of bacteria and plasmids promote genetic exchange^2,8,9^. Commensal bacteria inhabiting the human intestine, particularly in individuals living in low-resource settings, carry high levels of AMR genes and plasmids^10–13^. Vietnam has a high incidence of resistance to cephalosporins due to high consumption and easy access to these antibiotics^14,15^. Around 83% of healthy children in Vietnam carry ESBL-producing *Escherichia coli*^16^, while ESBL-producing *Enterobacterales* account for 60 and 90% of clinical samples^17–20^.

While metagenomic sequencing has highlighted human gut bacteria are a reservoir of AMR^21^, the extent of contribution by conjugative plasmids in these microbes to the accumulation of AMR in human pathogens remains poorly understood.

Traditionally, plasmids are classified into different incompatibility groups (Inc typing) based on their replicons^22^. Molecular surveillance has often relied on the low-resolution Inc typing method to detect plasmid transmission between lineages and species^23,24^. However, plasmids can evolve within and between bacterial species, forming populations with complex sub-structures that are not captured by standard Inc grouping^25–28^. Therefore, higher-resolution methods relying on sequence analysis, such as average nucleotide identity (ANI)^28,29^, Jaccard index (JI)^30,31^ and Mash distance^27,31^, have been developed to capture full genetic structure, plasmid relatedness and transmission events.

In this study, we utilised plasmid similarity network analyses combined with phylogenetic tools to investigate the structure and exchange of ESBL plasmids in the gut microbiome of healthy children, as well as putative plasmids in ceftriaxone-resistant *Escherichia coli* ST131 causing bloodstream infection, diarrhoeal *Shigella sonnei* and non-typhoidal *Salmonella* (NTS) and animal *E. coli*. Samples were collected within Vietnam over overlapping time periods.

## Methods

### Study samples

Samples were collected from a longitudinal cohort study monitoring diarrhoeal disease episodes in children in Ho Chi Minh City (HCMC), Vietnam. The study enrolled 748 children aged 12-36 months between June and December 2014 and followed them until December 2016 (OxTREC number: 1058-13)^32^. A total of 498 rectal swabs were collected at routine follow-up visits from children without a recent history of diarrhoeal disease^19^.

### Publicly available bacterial genomes from Vietnam

*S. sonnei* and NTS isolates were collected from children with diarrhoea in HCMC between 2014 and 2016 (OxTREC number: 0109)^13,33,34^. This dataset included raw Illumina reads of 47 ceftriaxone-resistant *S. sonnei* and 58 ceftriaxone-resistant NTS (Table S1).

Extra-intestinal pathogenic *E. coli* (ExPEC) isolates were collected from patients with bloodstream infection at Hospital for Tropical Diseases, HCMC between 2010 and 2014 (approval number: CS/ND/14/20)^35^. Raw Illumina reads of 86 ceftriaxone-resistant ExPEC ST131 isolates were included (Table S1).

*E. coli* isolates from faecal samples were obtained from chickens, pigs and farmers at different farms in Vietnam between 2015 and 2018^36,37^. Raw Illumina reads of 104 ceftriaxone-resistant *E. coli* isolates from chickens and pigs were included (Table S1).

### Isolation and sequencing of conjugative ESBL plasmids from healthy children

From 498 rectal swabs, 400 yielded ceftriaxone-resistant colonies after overnight culture. Sweeps of ceftriaxone-resistant colonies were pooled, and 254 pooled samples were randomly selected for ESBL plasmid isolation by bacterial conjugation with the recipient *E. coli* J53 (resistant to sodium azide and amikacin). After 24 hours of conjugation, transconjugants containing a conjugative ESBL plasmid were selected on MC agar supplemented with ceftriaxone (6 mg/L), sodium azide (100 mg/L) and amikacin (64 mg/L). From each individual, a single transconjugant was picked for plasmid extraction and plasmid DNA was sequenced using MiSeq Illumina.

### Sequence analysis of ESBL plasmids

Raw Illumina reads of ESBL plasmids were assembled using Unicycler v0.4.7^38^ and annotated with Bakta v1.5.0^39^. AMR genes and plasmid replicons were identified with ABRicate v1.0.0 (https://github.com/tseemann/abricate) against CARD^40^ and PlasmidFinder^41^ databases.

Whole genome sequences of *S. sonnei*, ExPEC ST131, NTS and animal *E. coli* were assembled and annotated using the same pipelines. Subsequently, ABACAS v1.3.1^42^ was used to map contigs against the following reference sequences: for *S. sonnei*, a concatenated sequence containing *S. sonnei* Ss046 (NC_007382), pSs046 virulence plasmid (NC_007385.1) and three small plasmids spA (NC_009345.1), spB (NC_009346.1) and spC (NC_009347.1); for NTS, *Salmonella* Typhimurium LT2 (AE006468.1); for ExPEC ST131 and animal *E. coli, E. coli* ST131 NCTC 13441 genome (NZ_LT632320.1). The unmapped contigs, presumably containing plasmids, were screened for AMR genes and plasmid replicons using ABRicate.

IncI plasmid Multi-Locus Sequence Typing (pMLST) was analysed using PubMLST (https://pubmlst.org).

### Phylogeny of IncI1 and IncB/O/K/Z plasmids

Raw Illumina reads from plasmids of healthy children and bacterial isolates containing IncI1 and IncB/O/K/Z were mapped to the reference IncI1 pKHSB.1 (NC_020991.1) and IncB/O/K/Z p02_1427 plasmid (ERR3094237) respectively, using Snippy v4.6.0 pipeline (https://github.com/tseemann/snippy). SNPs identified within repetitive regions were subsequently removed and the resulting SNP alignments (3594 SNPs for IncI and 5009 SNPs for IncB/O/K/Z) were used to infer maximum likelihood (ML) phylogenies with IQ-TREE v2.2.0.3^43,44^, incorporating best-fit substitution models and 1,000 ultrafast bootstraps.

For the global plasmid phylogeny, publicly available IncI or IncB/O/K/Z plasmids were downloaded from PLSDB database^45,46^, and screened for AMR genes using ABRicate^40^. *Bla*_*CMY*_*/bla*_*CTX-M*_-containing plasmids were combined with our dataset and mapped to the same reference IncI1 and IncB/O/K/Z as above. For SNP alignments, taxa with >50% gaps/ambiguities were removed, and only SNP columns with ≥80% completeness were retained. The resulting SNP alignments were used to infer ML phylogenies with IQ-TREE v2.2.0.3^43,44^.

### IncF plasmid analysis

Network analysis was performed to compare the structure of IncF plasmids, given their extensive diversity. A database of 54 complete IncF plasmid was compiled, comprising 24 from healthy gut microbiome, 5 from ExPEC ST131 isolates, and 25 reference Vietnamese plasmids from PLSDB database^45,46^ (Table S2), representing well-characterised ST131 IncF plasmids or highly similar plasmids as determined by *mash screen*^45,46^.

First, we calculated Jaccard index (JI) amongst the 54 plasmids using Mash v2.3^47^. JI between two plasmids *a* and *b* was calculated as

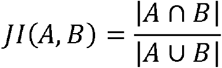

where *A* and *B* are the sets of *k*-mers of plasmids *a* and *b* respectively. This represents the extent of shared k-mers, where 0 means the two plasmids are dissimilar. Next, we computed containment index of the 54 plasmids within the mixture of unmapped contigs, presumably containing IncF plasmid sequences using Mash v2.3^48^. The containment index of plasmid *a* within the mixture of unmapped contigs *b* is calculated as

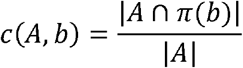

where *A* is the sets of *k*-mers of plasmid *a*, and π(*b*) is all of *b*. This represents the extent of k-mers in plasmid *a* (among the 54 plasmids) that was also found within the unmapped contigs of a bacterial isolate *b*, where 0 indicates no overlap.

Plasmid networks were visualised on Cytoscape v3.9.1^49^, with edges having JI or containment index < 0.75 removed, using a threshold higher than those previously reported^30,31^.

### Bacterial conjugation

We hypothesised that plasmid variants within distinct phylogenetic clusters may differ in conjugation frequencies. To test this, representative plasmid variants were purified from clinical isolates and introduced into *E. coli* MG1655 by electroporation. Conjugation was performed between Cro^R^ Tet^S^ *E. coli* MG1655 carrying different plasmid variants as the donor, and Cro^S^ Tet^R^ *E. coli* MG1655:zde264:Tn10 as the recipient^52^. After 4 hours of conjugation, donors, recipients and transconjugants were selected on MC plates with appropriate antibiotics. Conjugation frequency was calculated as the number of transconjugants per recipient^53^.

### Nanopore sequencing

Extracted plasmid or genomic DNA was subjected to Rapid Barcoding kit (Oxford Nanopore Technology-ONT), and sequenced on a Flongle flow cell (R9.4.1/R10.4.1). Basecalling and *de novo* assembly were performed using guppy v6.3.8 and Unicyler v0.4.7^38^ for plasmids, or dorado v0.8.0 (ONT), and Flye v2.9.5 for bacterial genomes.

## Results

### Distribution of ESBL plasmids in bacteria from different sources

Initially, we isolated conjugative ESBL plasmids from bacteria in the gut of healthy children as they play an important role in AMR spread, compared to non-conjugative or mobilisable plasmids^29^. Of 254 rectal swabs harbouring ceftriaxone-resistant bacteria, 77 samples (33.4%) contained conjugative ESBL plasmids. We successfully obtained sequences of 68 ESBL plasmids from these 77 samples.

IncF was the most prevalent plasmid group (42.7%, 29/68) isolated from the gut of healthy children, followed by IncB/O/K/Z (30.9%, 21/68), IncI1 (23.5%, 16/68) and IncX (2.9%, 2/68) (Figure 1). IncF plasmids carried mainly *bla*_CTX-M-27_ (70.0%, 20/29), followed by *bla*_CTX-M-14_ (13.8%, 4/29), *bla*_CTX-M-55_ (6.9%, 2/29) with one example of *bla*_CTX-M-15_ (3.4%). IncI1 plasmids were mostly associated with *bla*_CTX-M-15_ (62.5%, 10/16), followed by *bla*_CMY-42_ (18.75%, 3/16) and *bla*_CMY-44_ (18.75%, 3/16), while IncB/O/K/Z plasmids more frequently harboured *bla*_CTX-M-55_ (42.9%, 9/21) and bla_CTX-M-14_ (38.1%, 8/21).

**Figure 1:**
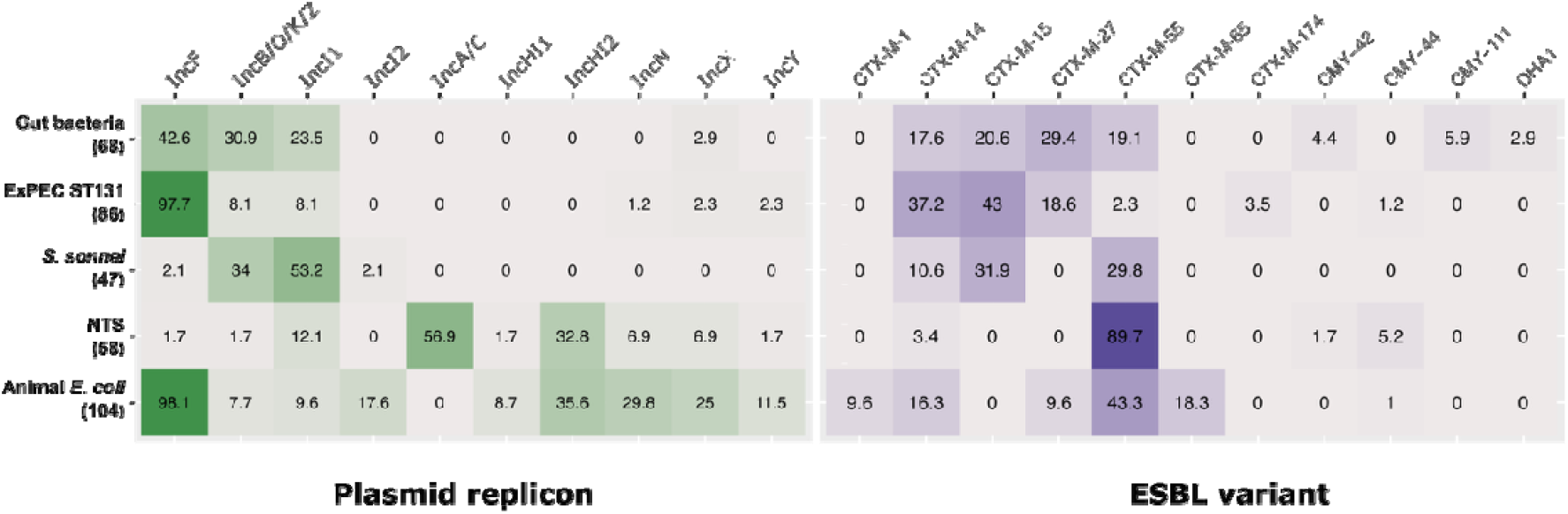
Plasmid replicon typing and ESBL characteristics of plasmids from the human gut bacteria, ExPEC ST131, diarrhoea-causing *S. sonnei* and non-typhoidal *Salmonella*, and *E. coli* from animals. Plasmid replicon types (green shading, left panel) and ESBL characteristics (purple shading, right panel) of plasmids from the human gut bacteria, ExPEC ST131, *S. sonnei* and NTS from patients with diarrhoea, and *E. coli* from animals. The percentage of Inc types and ESBL variants in isolates is given in the brackets.

Overall, the plasmid profiles and ESBL genes identified from healthy children were most similar to those observed in ExPEC ST131 and *S. sonnei*, characterised by the prevalence of IncF, IncI1 and IncB/O/K/Z plasmids, and the three main ESBL gene variants of *bla*_CTX-M-14_, *bla*_CTX-M-15_ and *bla*_CTX-M-55_ (Figure S1). Interestingly, *bla*_CTX-M-27_ was detected in both the human gut microbiome (29.4%) and ExPEC ST131 (18.6%) but not *S. sonnei*. In contrast, both NTS and animal *E. coli* isolates contained highly diverse yet similar patterns of plasmids (Figure 1). IncF plasmids dominated in animal *E. coli* (98.1%), while IncA/C plasmids (56.9%) were most prevalent in NTS. Notably, both NTS and animal *E. coli* carried IncHI2 (32.8% and 35.6%, respectively) and IncHI1 (1.7% and 8.7%, respectively) plasmids, which were not found in other bacterial hosts. Moreover, both NTS (89.7%) and animal *E. coli* (43.3%) isolates predominantly carried *bla*_CTX-M-55_. Notably, *bla*_CTX-M-65_ and *bla*_CTX-M-1_ were exclusively found in animal *E. coli*.

### Variants of IncB/O/K/Z plasmids proliferate via horizontal transfer or clonal expansion

We next investigated the phylogeny of IncB/O/K/Z plasmids identified in gut commensals from healthy children and other bacterial isolates (Figure 2A). The IncB/O/K/Z plasmids are divided into five major clusters (designated C1-5). C1, C2 and C4 plasmids displayed limited genetic diversity, with within-cluster median pairwise differences ranging from 1 to 3 SNPs (Table S3). In contrast, C3 and C5 plasmids displayed higher diversity, with median pairwise differences of 448 and 270 SNPs, respectively. *bla*_CTX-M-55_ was predominantly found in C1 and C2 plasmids, while *bla*_CTX-M-14_ was prevalent in C3 and C4 plasmids. C5 plasmids were associated with *bla*_CTX-M-65_, *bla*_CTX-M-55_ and bla_CTX-M-14_.

**Figure 2:**
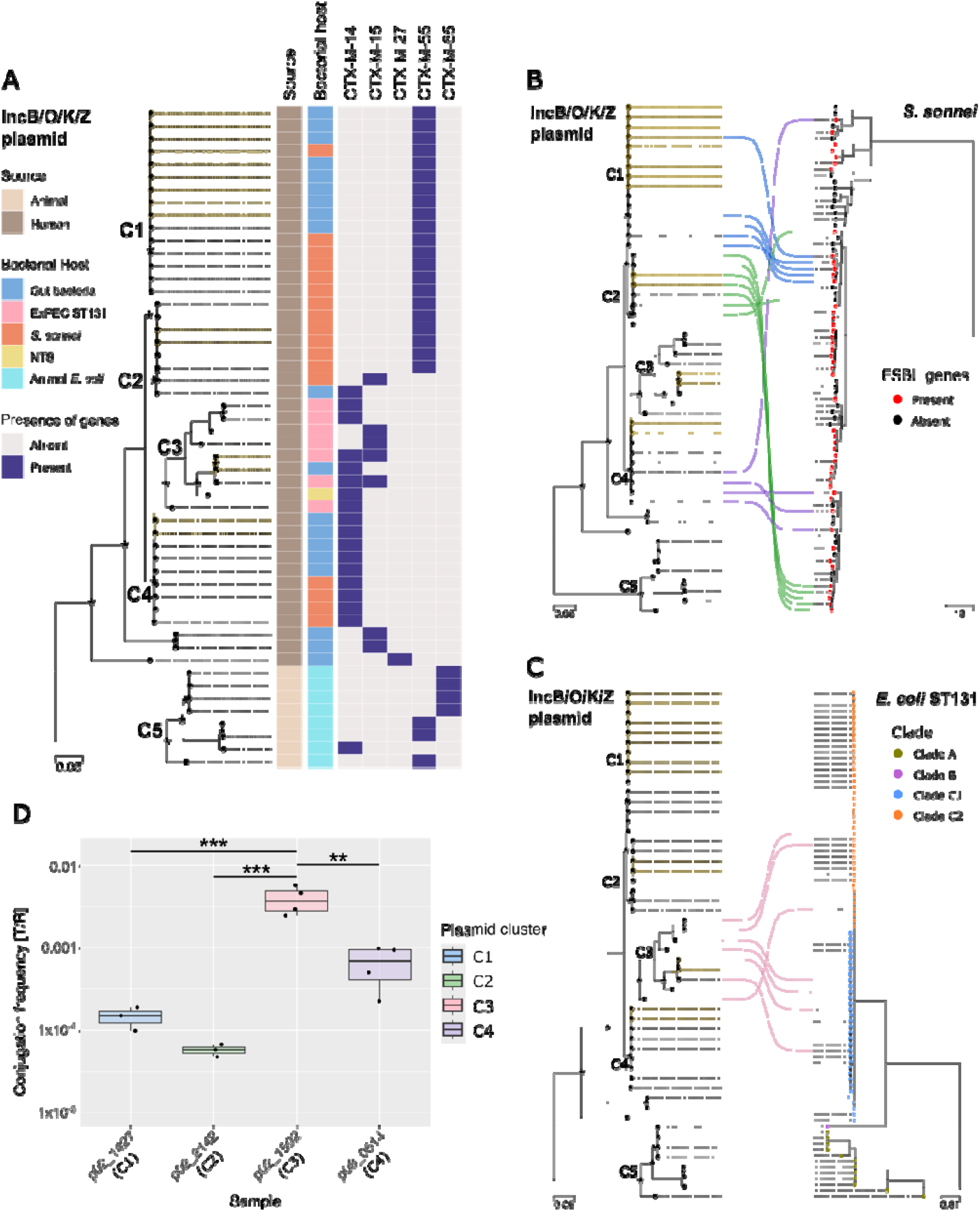
The landscape and characteristics of IncB/O/K/Z plasmids in Vietnam. **(A)** The phylogenetic structure of IncB/O/K/Z plasmids. The heatmap shows their source, bacterial host, and the presence (blue) or absence (grey) of different ESBL variants. The five clusters (C1, C2, C3, C4 and C5) of IncB/O/K/Z plasmids were identified. Identical plasmids are highlighted in yellow. The bar shows the number of substitutions per site. The black stars indicate nodes with bootstrap values higher than 85%. **(B)** The tanglegram links phylogenetic tree of IncB/O/K/Z plasmids (left) with the phylogenetic tree of *S. sonnei* isolates in Vietnam (right, modified from the previous study^13^). The node colours of *S. sonnei* phylogenetic tree (right) correspond to the absence or presence of ESBL genes. The lines connecting the trees are coloured by the clusters of IncB/O/K/Z plasmids. **(C)** The tanglegram links phylogenetic tree of IncB/O/K/Z plasmids (left) to the phylogenetic tree of ceftriaxone-resistant ExPEC ST131 isolates in Vietnam (right). The node colours of ExPEC ST131 phylogenetic tree (right) correspond to the ST131 clades. The lines connecting the trees are coloured by the clusters of IncB/O/K/Z plasmids. **(D)** The conjugation frequency of different ESBL-encoding IncB/O/K/Z plasmids in isogenic matings with *E. coli* MG1655 at 4h. IncB/O/K/Z plasmids were transferred from four different *S. sonnei* (02_1427, 02_2142 and 03_0514) and NTS strains (02_1592) to *E. coli* MG1655. Conjugation experiments were performed 3-4 times: the line shows the mean; the whiskers, the standard deviation. Plasmids belong to different clusters are indicated by different colours.

Notably, C1-C4 plasmids were detected from human sources (*i*.*e*., the gut microbiome and pathogenic bacteria), while C5 formed a separate lineage and were exclusively found in animal *E. coli*. C3 plasmids displayed the widest distribution across human gut bacteria, NTS, and ExPEC ST131, but not in *S. sonnei*. C1, C2 and C4 plasmids were detected both in the human microbiome and *S. sonnei*, although C2 plasmids were mostly found in *S. sonnei*. We also identified five identical plasmids, two of which were detected in both the human gut microbiome and either *S. sonnei* or ExPEC ST131, suggesting recent horizontal gene transfer (HGT) e1vents (Figure 2A).

We further assessed the transmission dynamics of IncB/O/K/Z plasmid by examining the congruence between plasmid and host phylogenies of *S. sonnei* (Figure 2B) and ExPEC ST131 isolates (Figure 2C). C1 plasmids were associated with a single *S. sonnei* cluster, consistent with vertical transmission. In contrast, C3 and C4 plasmids were linked to multiple acquisitions across distinct ExPEC ST131 and *S. sonnei* lineages, respectively, without evidence of clonal expansion, suggesting they disseminate predominantly *via* horizontal transfer. C2 plasmids was linked to three acquisition events in *S. sonnei*, one of which was associated with clonal spread.

The C3 plasmid, which possesses the broadest host range, exhibited the highest conjugation frequency (3.91 × 10^-3^, *p* < 0.05) (Figure 2D), followed by the C4 plasmid at a frequency of 6.6 × 10^-4^, and then the C1 plasmid at intermediate frequency of 1.46 × 10^-4^. In comparison, the C2 plasmid, which exhibits the lowest cross-species distribution, transferred at the lowest frequency (5.7 × 10^-5^). These data suggest a correlation between the plasmid transfer rate and their distribution in different clades and species.

### The dominant IncI1 plasmid variant displays effective horizontal transfer and clonal expansion

Next, we inferred a phylogenetic tree of IncI1 plasmids originating from the human gut bacteria and other bacterial isolates. The IncI1 plasmid phylogeny revealed three main lineages: lineage 1 and lineage 2 were marked by the expansion of three major plasmid groups, while for lineage 3 there were no instances of closely related plasmids (Figure 3A).

**Figure 3:**
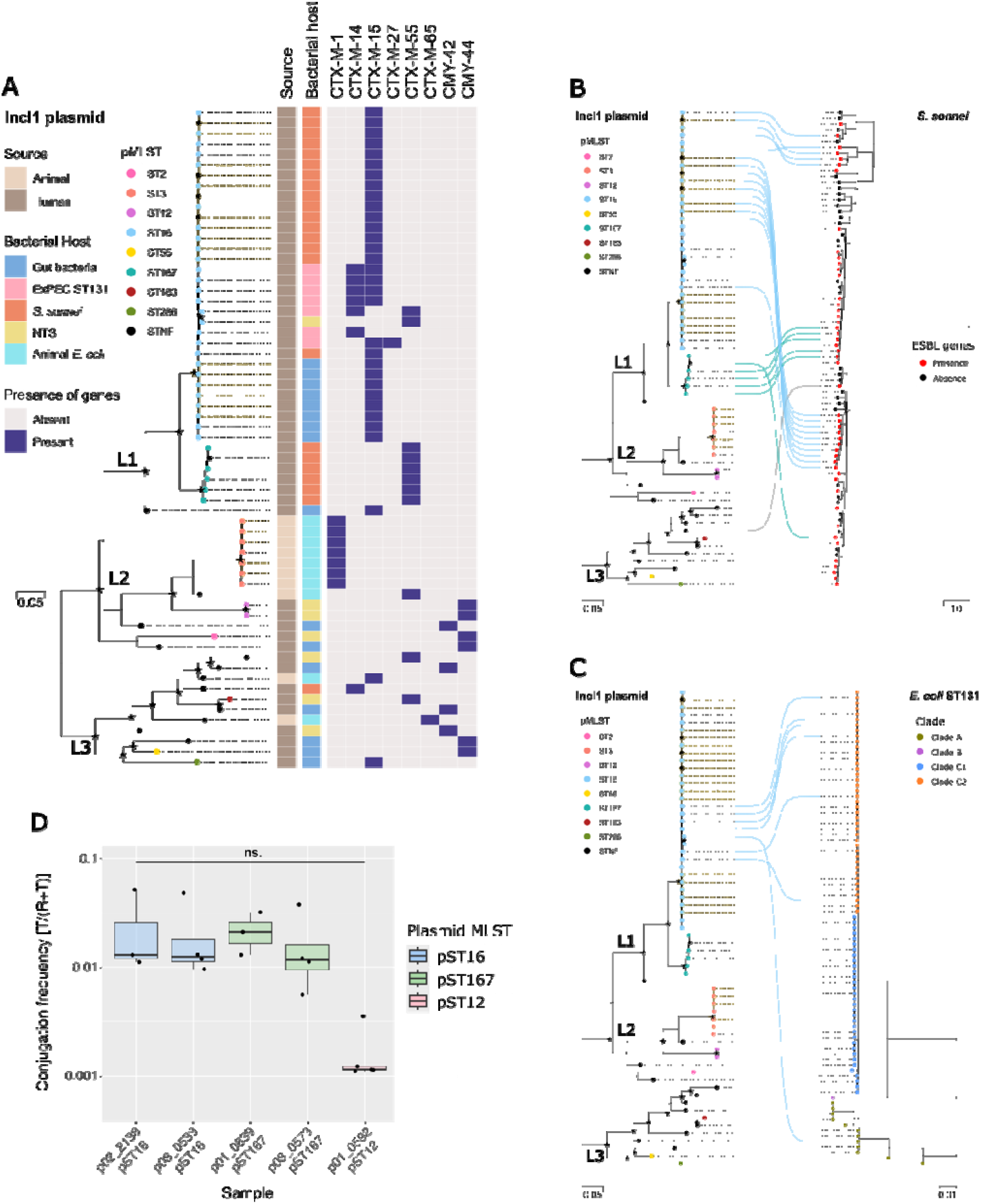
The landscape and characteristics of IncI1 plasmids in Vietnam. **(A)** The phylogenetic structure of IncI1 plasmids. The node colours correspond to different plasmid typing based on pMLST. The heatmap shows the source of isolation, bacterial host of the plasmids and the presence (blue) or absence (grey) of different ESBL variants. Identical plasmids are highlighted in yellow. The bar shows the number of substitutions per site. The black stars indicate nodes with bootstrap values higher than 85%. **(B)** The tanglegram links phylogenetic tree of IncI1 plasmids (left) to the phylogenetic tree of *S. sonnei* isolates in Vietnam (right, modified from the previous study^13^). The node colours of *S. sonnei* phylogenetic tree (right) correspond to the absence or presence of ESBL genes. The lines connecting the trees are coloured by the clusters of IncI1 plasmids. **(C)** The tanglegram links phylogenetic tree of IncI1 plasmids (left) to the phylogenetic tree of ceftriaxone-resistant ExPEC ST131 isolates in Vietnam (right). The node colours of ExPEC ST131 phylogenetic tree (right) correspond to the ST131 clades. The lines connecting the trees are coloured by the clusters of IncI1 plasmids. (D) The conjugation frequency of different ESBL-encoding IncI1 plasmids in *E. coli* MG1655 to Tet *E. coli* MG1655 at 4 hours. IncI1 plasmids were transferred from five different *S. sonnei* (01_0839, 02_2138, 03_0533 and 03_0573) and NTS strains (01_0592) to *E. coli* MG1655. Conjugation experiments were performed 3-4 times: the line shows the mean; the whiskers, the standard deviation. Plasmids belong to different pMLSTs are indicated by different colours.

L1/pST16/*bla*_CTX-M-15_ plasmids were the most prevalent in lineage 1, exhibiting minimal diversity (median 3 SNPs; Table S4), and were distributed widely in the human gut bacteria (8/32), ExPEC ST131 (7/32), *S. sonnei* (16/32) and NTS (1/32). These results indicate widespread dissemination of pST16 plasmid across diverse bacterial hosts. Integrating plasmid and host phylogenies of *S. sonnei* and ExPEC ST131 isolates supports both intraspecies transfer and clonal expansion pST16-carrying *S. sonnei* clusters (Figure 3B and 3C). These findings indicate that the success of the pST16 plasmid is driven by both horizontal transmission and clonal expansion. L1/pST167/*bla*_CTX-M-55_ plasmids were distantly related to L1/pST16/*bla*_CTX-M-15_ (mean 228 SNPs, Table S4) and restricted to *S. sonnei*. L1/pST167 plasmids also exhibited limited diversity and spread in *S. sonnei* via two independent acquisitions, one followed by clonal expansion (Figure 3B). Lineage 2 was marked by the expansion of pST3/*bla*_CTX-M-1_ plasmids in animal *E. coli*, and pST12/*bla*_CMY-44_ plasmids in NTS. Lineage 3 plasmids displayed higher genetic diversity, limited clonal expansion, and were found across bacterial species with diverse array of ESBL genes.

Notably, we observed a higher conjugation frequency for IncI1 compared with IncB/O/K/Z plasmids, indicating that IncI1 plasmids are likely more mobile (Figure 3D). L1/pST16 and L1/pST167 plasmids conjugated at high frequencies (1.67 × 10^-2^ to 2.53 × 10^-2^, respectively) compared to a L2/pST12 (1.63 × 10^-3^). We also generated *E. coli* MG1655 carrying an ESBL plasmid from lineage 3. However, this plasmid was not conjugative and lacked T4SS genes (Figure S2).

### Global phylogenies of IncB/O/K/Z and IncI1 plasmids

To place the IncB/O/K/Z ESBL plasmids circulating in Vietnam into a global context, we inferred a global IncB/O/K/Z plasmid phylogeny (Figure 4A). Overall, we identified three distinct plasmid lineages (L1-L3). The C5 plasmids from Vietnam were nested within the L1, while C3 plasmids were distributed across L2 and C1, C2 and C4 plasmids grouped within L3. In line with the Vietnam dataset, L1 plasmids were mainly isolated from animal samples, whereas L2 and L3 plasmids were primarily obtained from human samples. The IncB/O/K/Z plasmid variants with higher conjugation frequencies, including C3 (L2) and C4 plasmids (L3), possessed both broader host range and wider geographical spread (Figure 4A). Conversely, C1 and C2 plasmids (L3), showing the lowest conjugation frequencies, were mainly found in Vietnam and closely related bacterial species.

**Figure 4:**
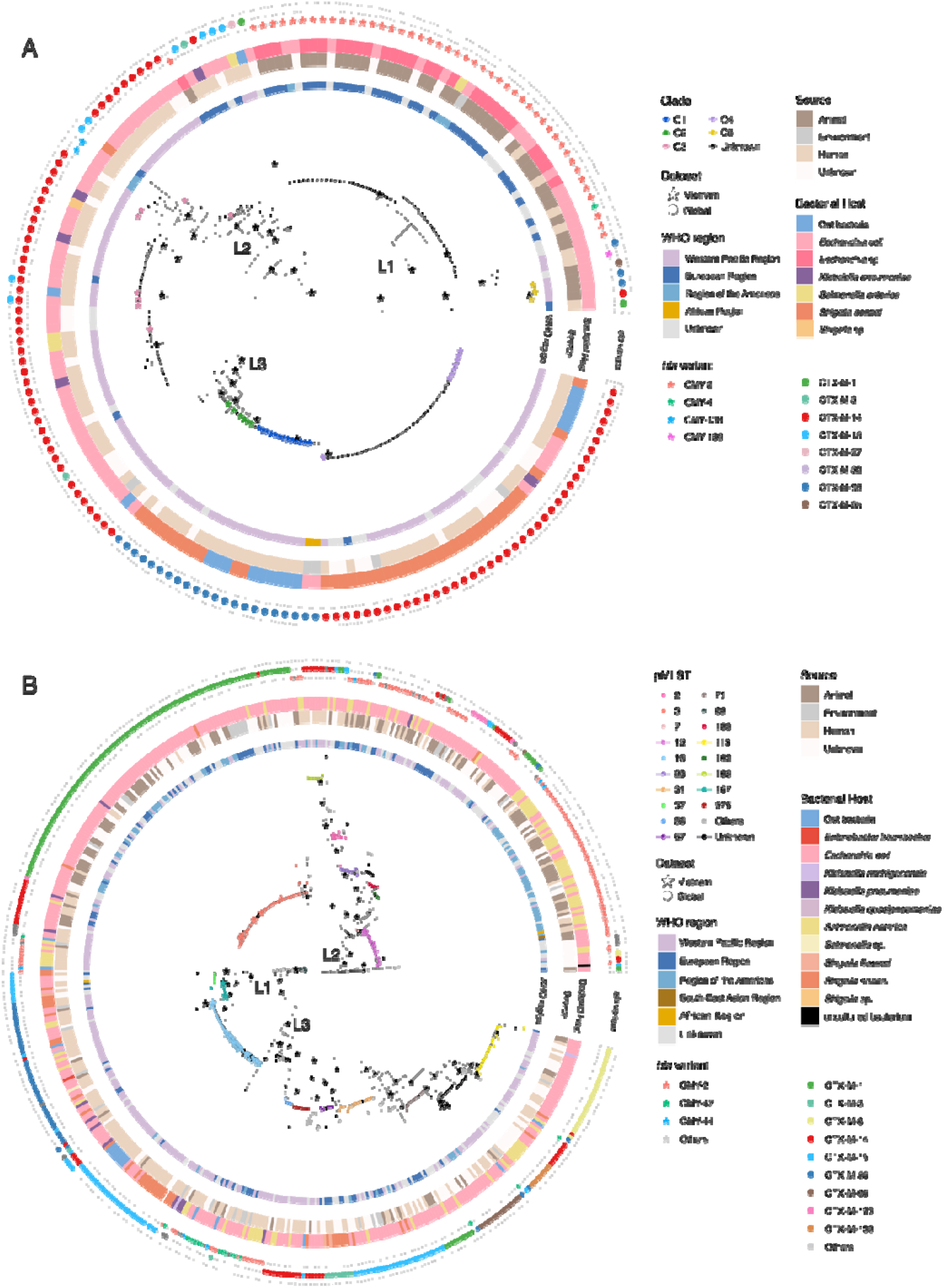
Global phylogeny of CMY- and CTX-M-carrying IncB/O/K/Z (A) and IncI1 (B) plasmids. The branches and tip points are coloured by either previously determined clusters (C1, C2, C3, C4 and C5) in **(A)** or pMLST in **(B)**. The tip labels show the ID of the plasmids and are coloured by WHO region. The heatmap shows the source and bacterial host. The stars show different CMY variants, while the circles show different CTX-M variants. The black stars indicate nodes with bootstrap values higher than 85%.

Similarly, a global IncI1 plasmid phylogeny revealed three lineages (L1-L3), consistent with the Vietnam dataset, with L1 and L3 plasmids found in human samples while L2 plasmids were largely found in animal samples (Figure 4B).

The most common lineage/pST/*bla*_CTX-M_ combinations were L2/pST3/*bla*_CTX-M-1_ (94/557), L1/pST16/*bla*_CTX-M-15_ or *bla*_CTX-M-55_ (63/557), and L2/pST12/*bla*_CMY-2_ (56/557). Both L2/pST3 and L2/pST12 displayed a restricted host range. Geographically, pST3 plasmids showed wide dispersal, whereas pST12 plasmids were largely confined to the Americas. L1/pST16 exhibited a broad host range and geographical dispersal, while L1/pST167 were predominantly associated with *S. sonnei, E. coli* and *S. enterica*, and sourced from the Western Pacific Region. Consistent with observations from IncB/O/K/Z, IncI variants with high conjugation frequencies (pST16 and pST167) were found across diverse hosts, whereas variant with lower conjugation frequency (pST12) was restricted to S. enterica or *E. coli*.

Collectively, the IncB/O/K/Z and IncI global plasmid phylogenies highlight a distinction between animal- and human-derived plasmids, with variants exhibiting higher conjugation frequencies typically associated with broader geographical and bacterial host distribution.

### Genetic structure and sharing of IncF plasmids

We identified ten clusters of IncF plasmids among the human gut bacteria and other bacterial isolates (Figure 5). Cluster 2 plasmids were the most prevalent (71/157), followed by cluster 1 (24/157) and cluster 3 (10/157). Cluster 2 plasmids shared a similar backbone with the pandemic F1:A2:B20/*bla*_CTX-M-14_ plasmid^25^ (Figure S3), but also carried other *bla*_CTX-M_ variants including *bla*_CTX-M-27_, *bla*_CTX-M-174_ and *bla*_CTX-M-15_. Notably, these plasmids were found across human gut microbiome, distinct clades of ExPEC ST131 (Figure 5D), and diverse STs in animal *E. coli*. These findings suggest that IncF cluster 2 plasmids can transfer within and between *E. coli* STs across human and animal. IncF cluster 1 plasmids were restricted to *bla*_CTX-M-15_-carrying ExPEC ST131 clade C2 (Figure 5D), while IncF cluster 3 plasmids carrying *bla*_CTX-M-55_ were mostly restricted to animal *E. coli* spanning across STs, with a single occurrence in ExPEC ST131.

**Figure 5:**
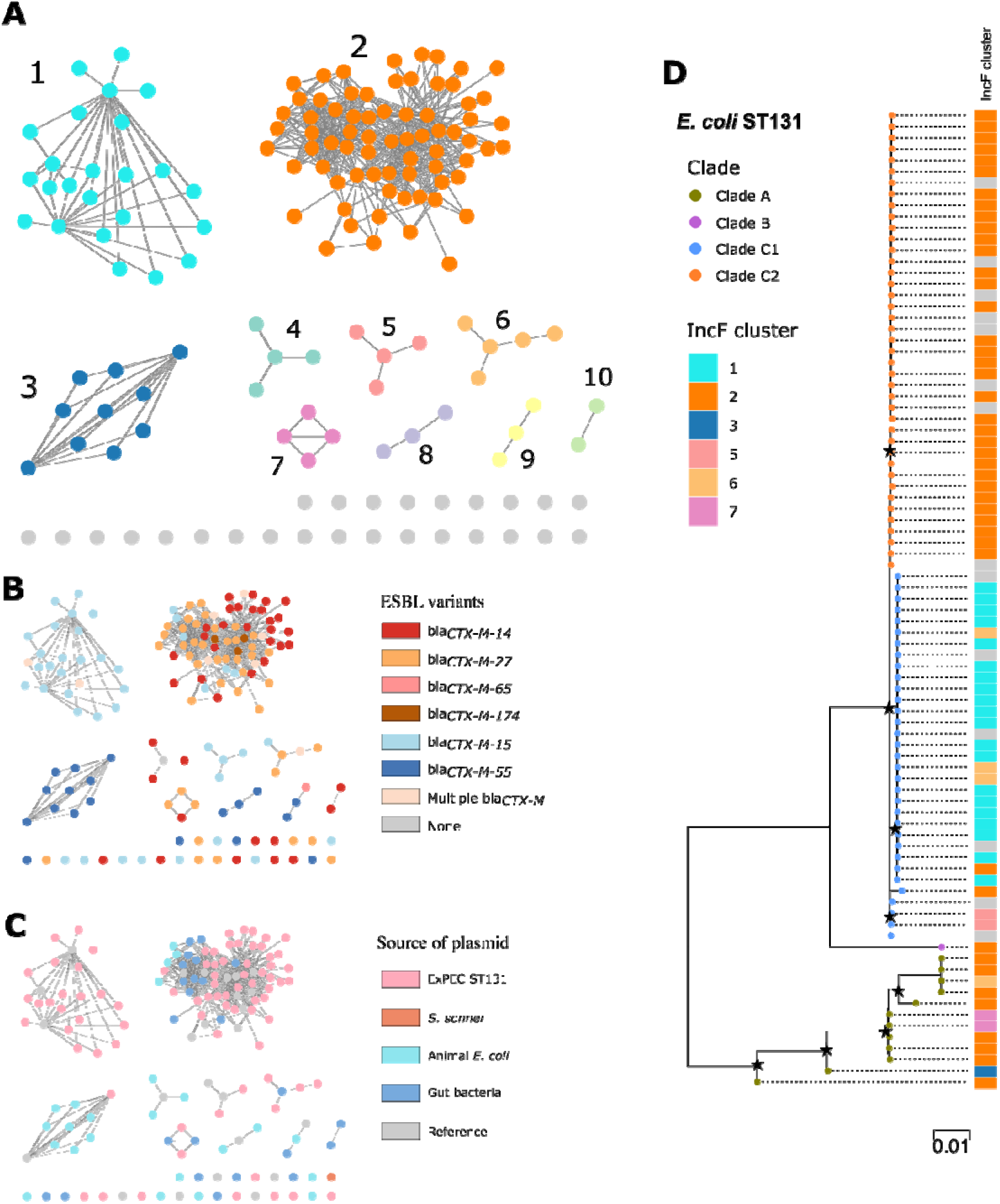
Plasmid similarity network of IncF plasmids at JI and containment index of 0.75. **(A)** Plasmid clusters are labelled from 1 to 10. Singletons are coloured grey. **(B)** Plasmid network coloured by different ESBL variants associated with either plasmids or bacterial isolates. **(C)** Plasmid network coloured by the source of isolation of either the plasmids or bacterial isolates. **(D)** Phylogenetic tree of ceftriaxone-resistant ExPEC ST131 isolates. The node colours correspond to the known ST131 clades. The heatmap shows the clusters of IncF plasmids associated to the ExPEC isolates. The black stars indicate nodes with bootstrap values higher than 85%.

## Discussion

Plasmids are the main vehicles for AMR dissemination, particularly in habitats with high bacterial density and diversity such as the human intestine^2,9,55^. However, there are limited data about the ecology and role of plasmids in the spread of ESBLs in low-resource settings. To address this gap, our study aimed to capture the prevalence, genetic structure, and transmission networks of common conjugative ESBL plasmids circulating in Vietnam.

We found high frequencies of self-transferable ESBL plasmids belonging to the IncF, IncI1 and IncB/O/K/Z groups in bacteria inhabiting the gastrointestinal tracts of healthy children. We also uncovered substantial evidence of these plasmids transferring between pathogens and human gut commensal bacteria, highlighting human gut microbiome as a key reservoir for ESBL plasmids for *S. sonnei*, ExPEC ST131 and, to a lesser extent, NTS. We also uncovered several prevalent plasmid/CTX-M combinations, including IncI1/pST16/*bla*_CTX-M-15_, IncB/O/K/Z/C1/*bla*_CTX-M-55_, IncB/O/K/Z/C4-*bla*_CTX-M-14_, IncF/C2/*bla*_CTX-M-14_ and IncF/C2-*bla*_CTX-M-_^27.^

By linking plasmid and chromosomal phylogenies with plasmid conjugation, our work elucidates the interplay between HGT for mediating spread and clonal expansion in the success of plasmids in bacterial populations. Amongst the IncI1 plasmids, the pST16 variant appeared to be the most successful, likely attributed its high conjugation frequency, and the tendency of pST16-carrying strains to undergo clonal expansion. In contrast, none of the IncB/O/K/Z variants illustrated such propensity. IncB/O/K/Z plasmid variants C1 and C2 with lower conjugation frequency were rarely associated with HGT; consequently, they were found in fewer bacterial populations but proliferated once moved into a bacterial host. Conversely, IncB/O/K/Z C3 and C4 variants, displaying higher conjugation frequencies, were associated with frequent transfer events but not clonal expansion, reflecting a potential lack of success of strains carrying these variants. These findings are in line with a previous model in which, at low plasmid cost, the effect of conjugation is negligible for plasmid persistence, but when plasmid fitness costs are substantial, high conjugation rates is vital for a plasmid being retained in a population^56^. Interestingly, from our global phylogenies, the highly conjugative plasmids such as IncI1/pST16, IncB/O/K/Z/C3 and IncB/O/K/Z/C4 were the most widespread plasmids, found across broad geographical and bacterial host range. Therefore, the plasmids most adept at achieving widespread epidemiological transmission are those that probably impose minimal costs on their host while retaining a high conjugation frequency.

The human microbiome is linked to the microbiome in domesticated animals and the surrounding environments^57,58^. Therefore, there has been increasing attention in tracking AMR dissemination between human, animal and environmental sources under a One Health approach^2^. Here, we observed little evidence of ESBL plasmid transmission between humans and animals in both Vietnam and global datasets. In fact, it appears that animal *E. coli* is likely a more significant AMR reservoir for plasmids in non-typhoidal *Salmonella*, a known coloniser of animal gut. Particularly, the genetic structure of ESBL plasmids recovered from animals was largely distinct from human-derived plasmids, especially for the IncB/O/K/Z and IncF plasmids. We also observed certain *bla*_CTX-M_ genes (*bla*_CTX-M-1_ and *bla*_CTX-M-65_) and plasmid groups (IncI1/pST3, IncB/O/K/Z/C5 and IncF/C3) were only detected in animal *E. coli*. Similarly, previous studies in Vietnam have shown limited AMR transmission between bacteria in humans and animals^37,59–61^. Differential diet, housing, lifestyle and genetics of humans and animals have shaped the divergent compositions of their microbiomes^62–65^. While domestication and captivity are associated with bacterial transmission from humans to animals^66–68^, there are a limited number of strains capable of colonising both hosts, with infrequent bacterial sharing^69^. Therefore, we speculate that plasmid sharing between these ecological niches is very infrequent and cannot be captured without a targeted approach. Accordingly, future One Health research may adopt a plasmid-centric framework that helps mitigate sampling bias and undersampling by enabling a more targeted tracking of cross-host transmission of clinically relevant AMR plasmids.

## Conclusions

Our study provides insights into the prevalence and genetic structure of three major groups of conjugative ESBL plasmids circulating in human gut microbiome in Vietnam, and how these plasmids are shared between human pathogens and animal *E. coli*. Importantly, we have identified epidemic plasmid variants whose dissemination is likely driven by both HGT and bacterial clonal expansion. Future studies will focus on molecular mechanisms responsible for their dissemination and costs of this process for a range of bacterial hosts. This would potentially inform the development of novel interventions to block plasmid transfer or promote plasmid loss as a means to slow down the global spread of AMR.

## Data Availability

All data used in this work are publicly available in the European Nucleotide Archive (ENA) under project numbers PRJEB96742 (https://www.ebi.ac.uk/ena/browser/view/PRJEB96742)

## Declarations

### Ethics approval and consent to participate

Samples used in this study were collected from a longitudinal cohort study monitoring diarrhoeal disease episodes in children in Ho Chi Minh City (HCMC), Vietnam. Ethical approval for the study was granted by Oxford Tropical Research Ethics Committee (OxTREC number: 1058-13)^32^. Informed consent was obtained from all participants prior to enrolment. All procedures were conducted in accordance with the Declaration of Helsinki and relevant institutional guidelines.

### Consent for publication

Not applicable

## Acknowledgements

We would like to thank the healthcare staff at the Hospital for Tropical Diseases, Ho Chi Minh City, Viet Nam for their assistance in data collection for this study.

## Author Contributions

P.P. collected, analysed and interpreted data, and conducted experiments under the supervision of P.T.D. and C.M.T. P.P. drafted and revised the manuscript with P.T.D. and C.M.T. P.T.M.Q. conducted experiments and contributed to data interpretation. L.T.Q.N., V.T.D., N.P.H.L, S.B., H.T.T. collected data and helped with manuscript revision. Q.N., C.T.H., N.T.N.T supported data collection, analysis and manuscript revision. M.A.R. and G.T. contributed to manuscript writing and revision. P.P., P.T.D. and C.M.T. conceptualised and designed the study, led the analysis and data interpretation and finalised the manuscript.

