## Supplementary figures for "Ecological and molecular drivers of ESBL plasmid dissemination in *Enterobacteriaceae* in Vietnam"

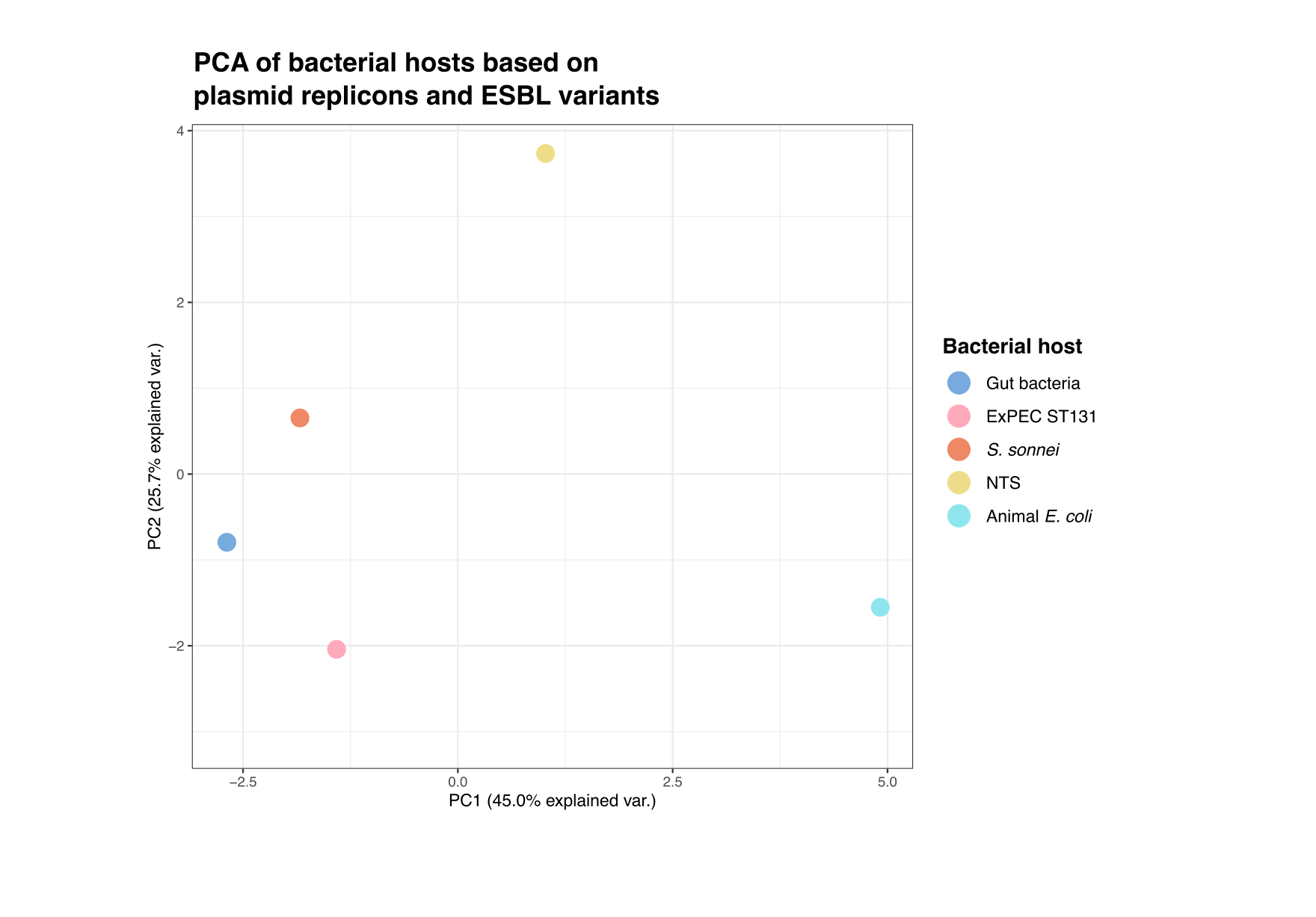

**Figure S1: Principal component analysis of bacterial hosts based on plasmid replicons and ESBL variants**

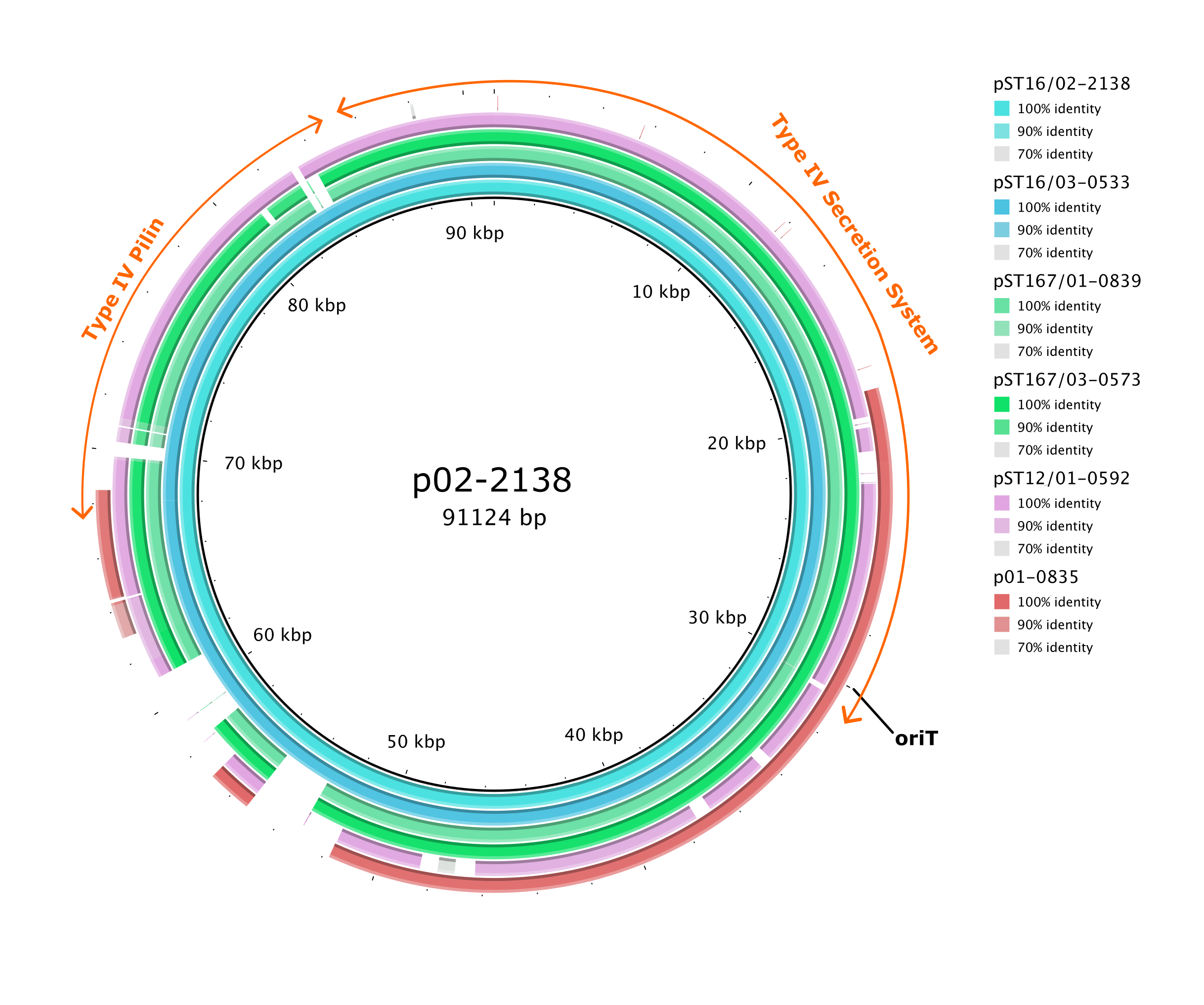

**Figure S2: BLASTN comparisons of IncI1 plasmids, using p02-2138 plasmid as the reference.**

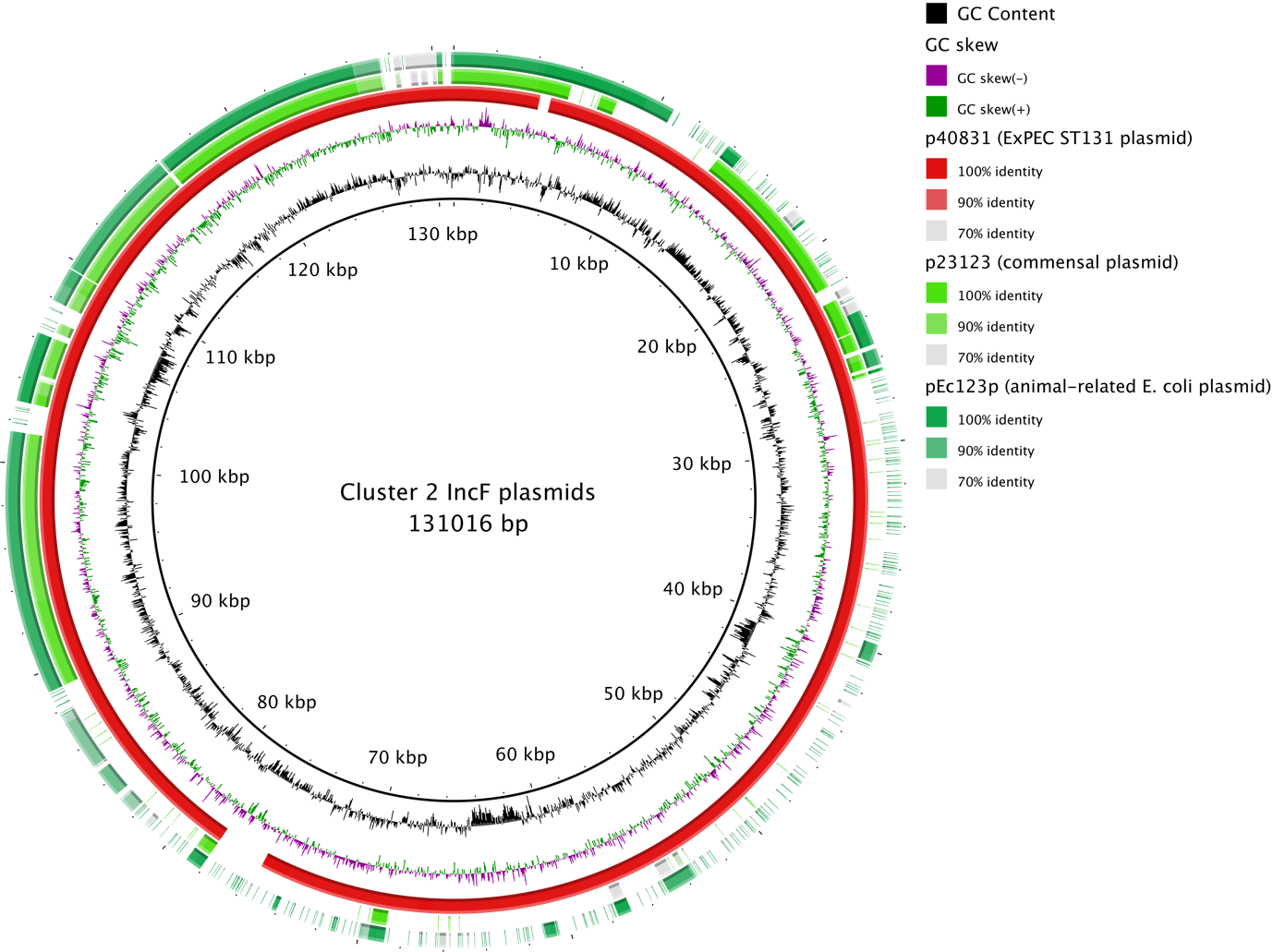

**Figure S3: BLASTN comparisons of cluster 2 IncF plasmids from ExPEC ST131, human gut bacteria and animal *E. coli*, using the pandemic F1:A2:B20 plasmid pMO (MG886288.1) as the reference (the central black circle).**
